# A serological assay to detect SARS-CoV-2 seroconversion in humans

**DOI:** 10.1101/2020.03.17.20037713

**Authors:** Fatima Amanat, Daniel Stadlbauer, Shirin Strohmeier, Thi H.O. Nguyen, Veronika Chromikova, Meagan McMahon, Kaijun Jiang, Guha Asthagiri Arunkumar, Denise Jurczyszak, Jose Polanco, Maria Bermudez-Gonzalez, Giulio Kleiner, Teresa Aydillo, Lisa Miorin, Daniel Fierer, Luz Amarilis Lugo, Erna Milunka Kojic, Jonathan Stoever, Sean T. H. Liu, Charlotte Cunningham-Rundles, Philip L. Felgner, Thomas Moran, Adolfo Garcia-Sastre, Daniel Caplivski, Allen Cheng, Katherine Kedzierska, Olli Vapalahti, Jussi M. Hepojoki, Viviana Simon, Florian Krammer

**Affiliations:** Department of Microbiology, Icahn School of Medicine at Mount Sinai, New York, NY, USA; Graduate School of Biomedical Sciences, Icahn School of Medicine at Mount Sinai, New York, NY, USA; Department of Biotechnology, University of Natural Resources and Life Sciences, Vienna, Austria; Department of Microbiology & Immunology, University of Melbourne, The Peter Doherty Institute for Infection & Immunity, Melbourne, Victoria, Australia; Department of Genetics and Genomic Sciences, Icahn School of Medicine at Mount Sinai, New York, NY, USA; Division of Infectious Diseases, Department of Medicine, Icahn School of Medicine at Mount Sinai, New York, NY, USA; Division of Pulmonary, Critical Care, and Sleep Medicine, Icahn School of Medicine at Mount Sinai, New York, NY, USA; Department of Pediatrics and Medicine, Icahn School of Medicine at Mount Sinai, New York, NY, USA; Department of Physiology & Biophysics, University of California, Irvine, CA, USA; Global Health Emerging Pathogens Institute, Icahn School of Medicine at Mount Sinai, NY, USA; Travel Medicine Program, Division of Infectious Diseases, Icahn School of Medicine at Mount Sinai, New York, NY, USA; School of Public Health and Preventive Medicine, Monash University; Infection Prevention and Healthcare Epidemiology Unit, Alfred Health; Department of Virology, Medicum, University of Helsinki, Helsinki, Finland; Veterinary Biosciences, Veterinary Faculty, University of Helsinki, Helsinki, Finland; Department of Virology and Immunology, Helsinki University Hospital (HUSLAB), Helsinki, Finland; Institute of Veterinary Pathology, Vetsuisse Faculty, University of Zürich, Zürich, Switzerland

## Abstract

SARS-Cov-2 (severe acute respiratory disease coronavirus 2), which causes Coronavirus Disease 2019 (COVID19) was first detected in China in late 2019 and has since then caused a global pandemic. While molecular assays to directly detect the viral genetic material are available for the diagnosis of acute infection, we currently lack serological assays suitable to specifically detect SARS-CoV-2 antibodies. Here we describe serological enzyme-linked immunosorbent assays (ELISA) that we developed using recombinant antigens derived from the spike protein of SARS-CoV-2. Using negative control samples representing pre-COVID 19 background immunity in the general adult population as well as samples from COVID19 patients, we demonstrate that these assays are sensitive and specific, allowing for screening and identification of COVID19 seroconverters using human plasma/serum as early as two days post COVID19 symptoms onset. Importantly, these assays do not require handling of infectious virus, can be adjusted to detect different antibody types and are amendable to scaling. Such serological assays are of critical importance to determine seroprevalence in a given population, define previous exposure and identify highly reactive human donors for the generation of convalescent serum as therapeutic. Sensitive and specific identification of coronavirus SARS-Cov-2 antibody titers may, in the future, also support screening of health care workers to identify those who are already immune and can be deployed to care for infected patients minimizing the risk of viral spread to colleagues and other patients.

## Introduction

On December 31^st^, 2019 China reported first cases of atypical pneumonia in Wuhan, the capital of Hubei province. The causative virus was found to be a betacoronavirus, closely related to the severe acute respiratory syndrome coronavirus (SARS-CoV-1) from 2003 and similar to *Sarbecoviruses* isolated from bats.^1,2^ It was therefore termed SARS-CoV-2 and the disease it causes was named COVID19 (COronaVIrus Disease 2019).^3^ The outbreak in Wuhan expanded quickly and led to the lockdown of Wuhan, the Hubei province and other parts of China. While the lockdown, at least temporarily, brought the situation under control in China, SARS-CoV-2 spread globally causing a pandemic with, so far, 1,700,000 infections and 107,000 fatalities (as of April 11^th^, 2020).

Nucleic acid tests that detect the SARS-CoV-2 RNA genome were quickly developed and are now widely employed to diagnose COVID19 disease.^4,5^ However, there remains a great need for laboratory assays that measure antibody responses and determine seroconversion. While such serological assays are not well suited to detect acute infections, they support a number of highly relevant applications. First, serological assays allow us to study the immune response(s) to SARS-CoV-2 in a qualitative and quantitative manner. Second, serosurveys are needed to determine the precise rate of infection in an affected area, which is an essential variable to accurately determine the infection fatality rate. Third, serological assays will allow for the identification of individuals who mounted strong antibody responses and who could serve as donors for the generation of convalescent serum therapeutics. Lastly, serological assays will potentially permit to determine who is immune and who is not. This information may be very useful for deploying immune healthcare workers in a strategic manner as to limit the risk of exposure and inadvertent spread of the virus. It could also allow a certain proportion of the population that has already acquired immunity to go back to ‘normal life’. Of course, parallel studies that determine which antibody titers correlate with protection are needed to support these potential measures.

*Sarbecoviruses* express a large (approximately 140 kDa) glycoprotein termed spike protein (S, a homotrimer), which mediates binding to host cells via interactions with the human receptor angiotensin converting enzyme 2 (ACE2).^6-8^ The S protein is very immunogenic with the receptor-binding domain (RBD) being the target of many neutralizing antibodies.^9^ Individuals infected with coronaviruses typically mount neutralizing antibodies, which might be associated with some level of protection for a period of months to years^10-12^, and a neutralizing response has been demonstrated for SARS-CoV-2 in an individual case from day 9 onwards.^13^ Serum neutralization can be measured using replication competent virus but the process requires several days and must be conducted in biosafety level 3 laboratory containment for SARS-CoV-2. Potentially, pseudotyped viral particle based entry assays using lentiviruses or vesicular stomatitis virus could be used but these reagents are not trivial to produce. A simple solution is the use of a binding assay, e.g. an enzyme linked immunosorbent assays (ELISA), with recombinant antigen as substrate. Here we report the development of such an assay and provide a protocol for both recombinant antigen production as well as the ELISA methodology.^14^ An assay based on our protocol was implemented at Mount Sinai’s clinical laboratory, has received emergency use authorization from New York State and is used for screening plasma donors with high titers of antibodies to SARS-CoV-2. We have also distributed our protocol and reagents to well above 200 laboratories across the globe.

## Results

### Expression constructs and generation of recombinant SARS-CoV-2 proteins

We generated two different versions of the SARS-CoV-2 spike protein. The first construct expresses a full length trimeric and stabilized version of the spike protein and the second one produces only the much smaller receptor binding domain (RBD). The sequence used for both proteins is based on the genomic sequence of the first virus isolate, Wuhan-Hu-1, which was released on January 10^th^ 2020.^1^ Sequences were codon optimized for mammalian cell expression. The full-length spike protein sequence was modified to remove the polybasic cleavage site, which is recognized by furin and to add a pair of stabilizing mutations (**Figure 1**).^7,15,16^ These two modifications were included to enhance the stability of the protein based on published literature.^7,15^ At amino acid P1213 the sequence was fused to a thrombin cleavage site, a T4 foldon sequence for proper trimerization and a C-terminal hexahistidine tag for purification (**Figure 1**).^17,18^ The sequence was cloned into a pCAGGS vector for expression in mammalian cells and into a modified pFastBac Dual vector for generation of baculoviruses and expression in insect cells. For expression of the RBD, the natural N-terminal signal peptide of S was fused to the RBD sequence (amino acid 319 to 541) and joined with a C-terminal hexahistidine tag.^19^ The same vectors as for the full length S protein were used to express the RBD. In mammalian cells, the RBD domain gave outstanding yields (approximately 25-50 mg/liter culture), but expression was lower in insect cells (approximately 1.5 mg/liter culture). Clear single bands were visible when the recombinant RBD proteins were analyzed on a reducing sodium dodecyl sulfate–polyacrylamide gel electrophoresis (SDS-PAGE), with the insect cell derived protein (iRBD) running slightly lower than the mammalian cell derived protein (mRBD) (**Figure 1**). The size difference likely reflects differences in glycan sizes between insect cells and mammalian cells. The full-length S protein was also expressed in both systems with slightly higher yields in mammalian cells (mSpike) than in insect cells (iSpike) (approximately 5 mg/liter cultures versus 0.5 mg/liter culture). The full-length protein appeared as prominent band between 135 and 190 kDa followed by a faint, second band slightly below on a reducing SDS-PAGE, the higher species likely being the full-length protein and the slightly lower species likely a cleavage product.

**Figure 1:**
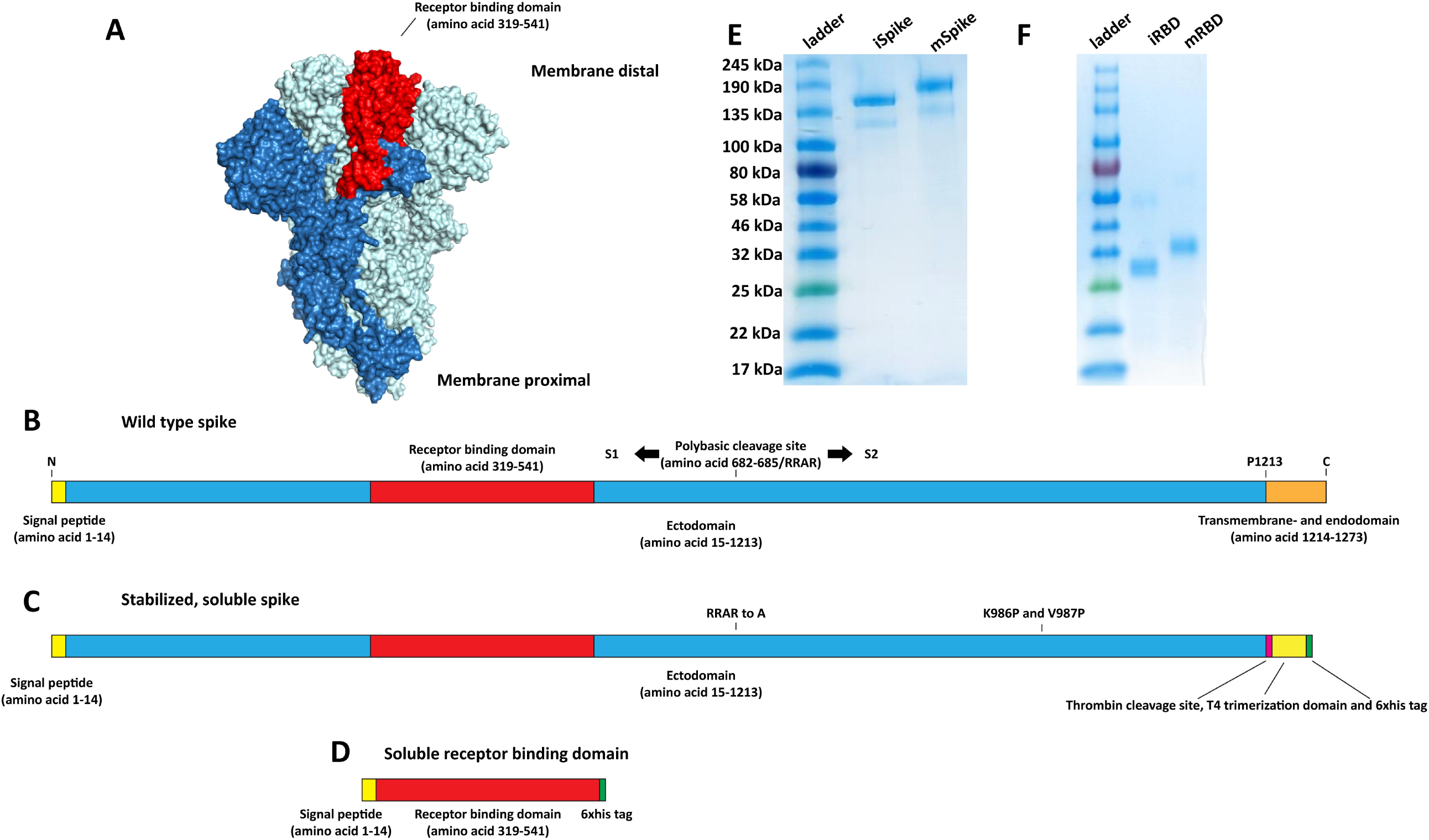
Constructs for recombinant protein expression. **A** Visualization of the trimeric spike protein of SARS-CoV-2 based on PBD # 6VXX using Pymol.^8^ One monomer is colored in dark blue while the remaining two monomers are held in light blue. The receptor binding domain (RBD) of the dark blue trimer is highlighted in red. **B** Schematic of the wild type full length spike protein with signal peptide, ectodomain, receptor binding domain, furin cleavage site, S1, S2, and transmembrane and endodomain domain indicated. **C** Schematic of the soluble trimeric spike. The polybasic/furin cleavage site (RRAR) was replaced by a single A. The transmembrane and endodomain were replaced by a furin cleavage site, a T4 foldon tetramerization domain and a hexahistidine tag. Introduction of K986P and V987P has been shown to stabilize the trimer in the pre-fusion conformation. **D** Schematic of the soluble receptor binding domain construct. All constructs are to scale. **E** Reducing SDS PAGE of insect cell and mammalian cell derived soluble trimerized spike protein (iSpike and mSpike). **F** Reducing SDS PAGE of insect cell derived and mammalian cell derived recombinant receptor binding domain (iRBD and mRBD).

### ELISA development

Initially, we tested a panel of 50 (59 for mRBD) banked human serum samples collected from study participants without and with confirmed previous viral infections (e.g., hantavirus, dengue virus, coronavirus NL63 – sample take 30 days post symptom onset) to establish an ELISA with our proteins. These human sera were used to test the background reactivity to the SARS-CoV-2 spike in the general US population covering an age range from approximately 20 to 65+ years. An initial set of four plasma/serum samples from three COVID19 survivors were used to determine the reactivity of SARS-CoV-2 infected individuals to the RBD and the full length spike (**Figure 2)**.

**Figure 2:**
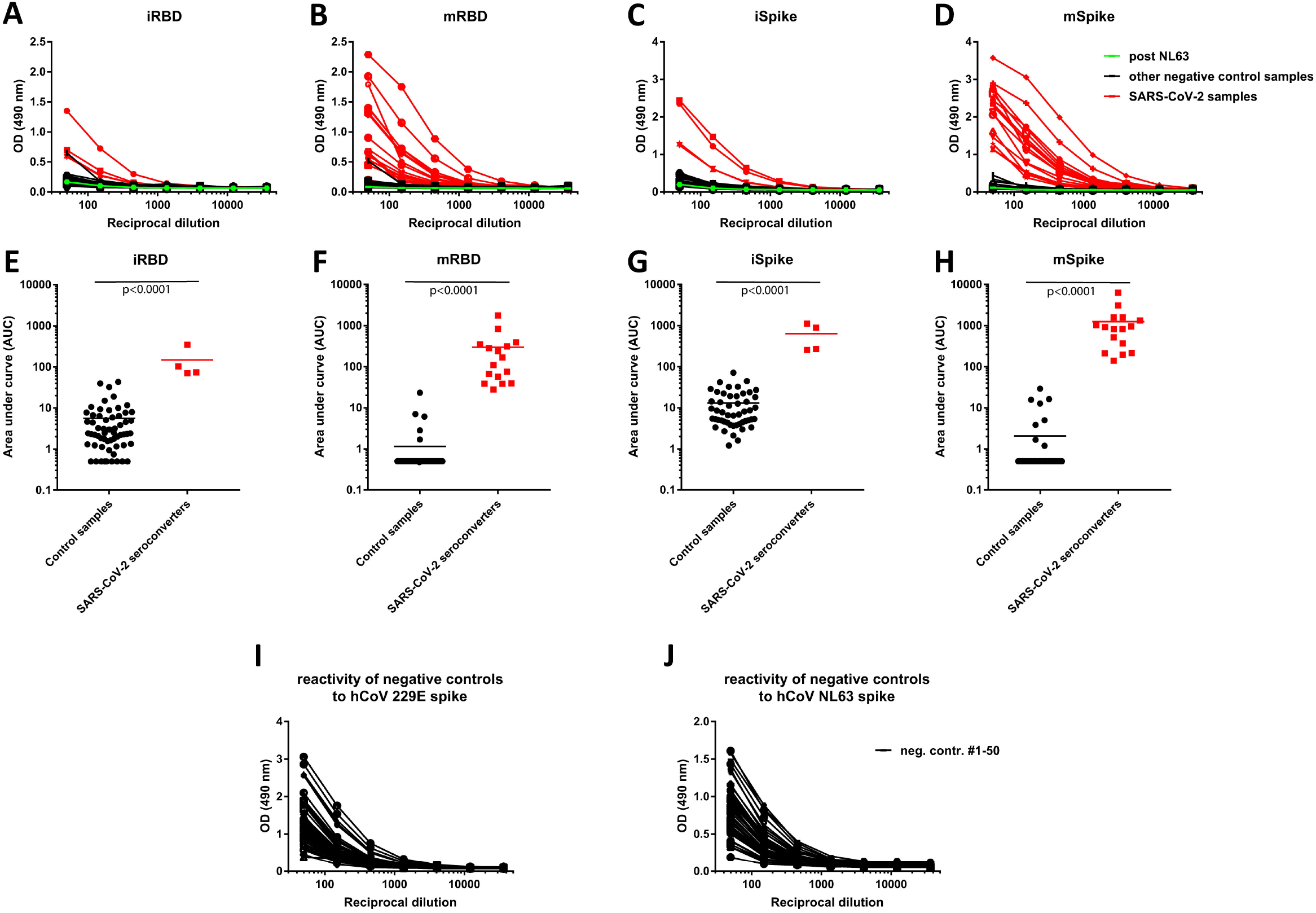
Reactivity of control and SARS-CoV-2 convalescent sera to different spike antigens. **A-D** Reactivity to insect cell derived RBD (iRBD), mammalian cell derived RBD (mRBD), insect cell derived soluble spike protein (iSpike) and mammalian cell derived soluble spike protein (mSpike). Sera from SARS-CoV-2 infected individuals are shown in red. One sample, shown in green, is a convalescent serum sample post NL63 infection. **E-F** shows data from the same experiment but graphed as area under the curve (AUC) to get a better quantitative impression. The n for the control samples is 50 except for the iRBD where it is 59. Statistics were performed using an unpaired two-tailed student’s t-test in Graphpad Prism. **I-G** shows reactivity of the 50 negative control samples from **A-F** against spike protein from human coronaviruses 229E and NL63.

ELISAs were performed by doing serial dilution of the individual serum samples. Values from the dilution curves were used to determine the area under the curve (AUC), which was graphed. All COVID19 plasma/serum samples reacted strongly to both RBD and full-length spike protein while reactivity of the other serum samples only yielded background reactivity (**Figure 2**). Reactivity of COVID19 sera was, in general, stronger against the full-length S protein than against the RBD, likely reflecting the higher number of epitopes found on the much larger spike protein. For the RBD the difference between control sera and convalescent sera was larger when the mammalian cell derived protein was used as compared to the insect cell derived RBD. The same was true for the full-length spike protein. Due to the expanding epidemic in New York City, we tested next an additional 14 serum samples from patients with acute COVID19 disease as well as convalescent participants for reactivity to mRBD and mSpike. These additional data were added to **Figure 2B, 2D, 2F and 2H**. All 14 samples reacted well with both RBD and spike protein. Thus, our assays allowed to clearly distinguish the sera from participants diagnosed with COVID19 from those collected prior to the pandemic (e.g., collected in the fall of 2019).

Our initial set of negative controls included convalescent serum from a participant with a confirmed NL63 infection. Importantly, this sample did not produce a signal against the SARS-CoV-2 RBD or spike underscoring the specificity of our assays. Since human coronaviruses OC43, 229E, NL63 and/or HKU1 are responsible for a large proportion of common colds every year, cross-reactivity between SARS-CoV-2 and these seasonal coronaviruses is of particular importance and warranted further investigation. To test how common antibodies to human corona viruses other than SARS-CoV are in our “pre-pandemic serum panel”, we performed ELISAs coated with spike protein of coronavirus 229E and NL63. While none of the negative control sera reacted to SARS-CoV-2 RBD and spike, the majority of samples yielded strong signals to the spike proteins of these two human coronaviruses **(Figure 2I and 2J**). In addition, we tested 21 different batches (27 vials) of pools of different products of normal human immune globulin (NHIG), intended for intravenous use and derived from >1000 donors each. None of the NHIG preparations reacted with SARS-CoV-2 RBD or spike protein and the signal obtained was similar to that of the three irrelevant human monoclonal antibodies (mAbs). The RBD-binding mAb CR3022 produced, in contrast, a strong signal **(Figure 3A and 3B**).^20-22^ Lastly, we tested panel of fifty plasma samples collected from HIV positive patients banked 2008 and 2011. Again, none of the samples reacted with the SARS-CoV-2 RBD or spike (**Figure 3C and 3D)**.

**Figure 3:**
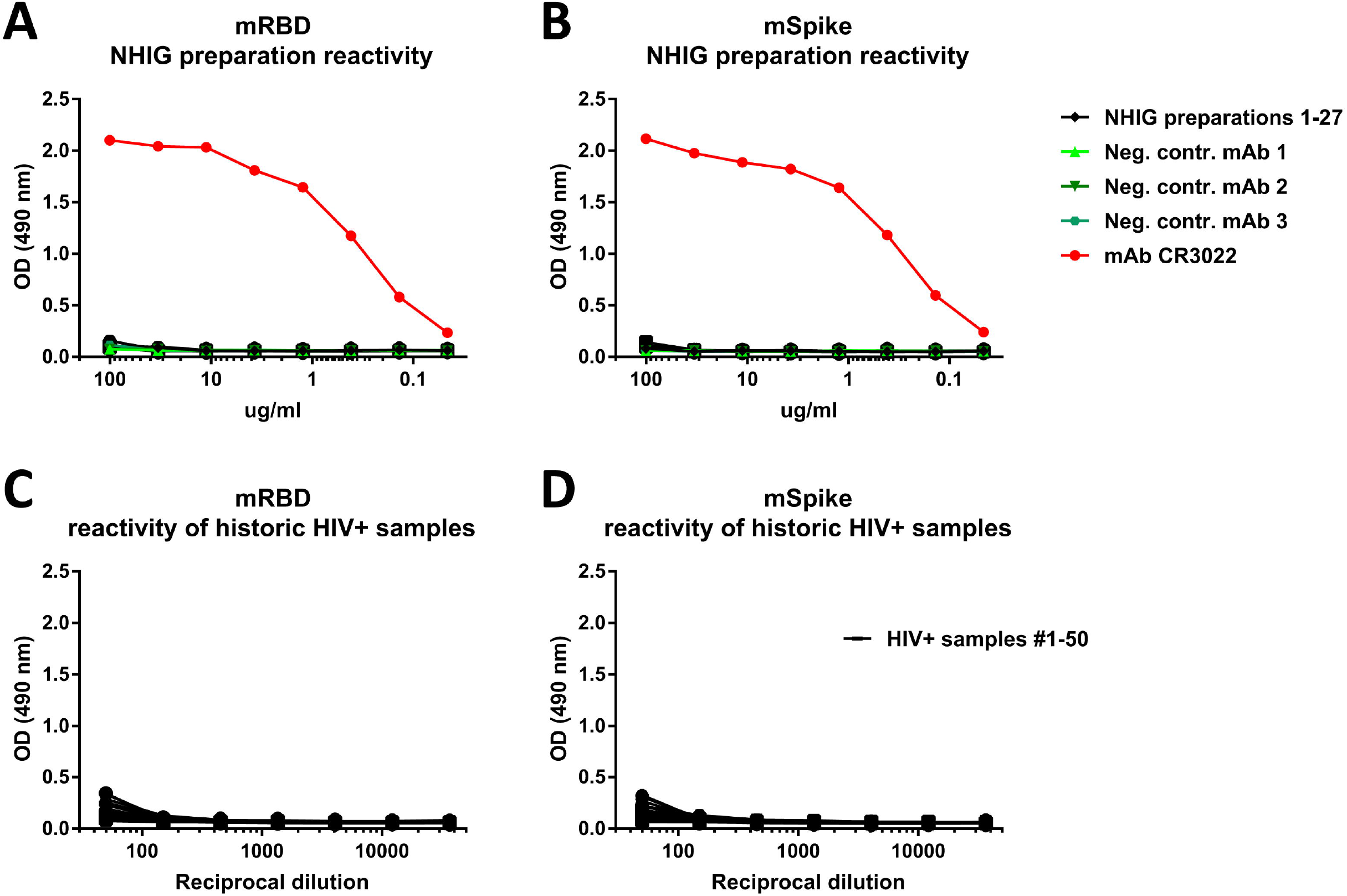
Figure 3: Human normal immunoglobulin preparations and historic sera from HIV+ patients do not react with the SAR-CoV-2 spike. **A-B** Reactivity of 21 different pools of human normal immunoglobulin (HNIG) preparations (27 different vials) to mRBD and mSpike of SARS-CoV-2. MAb CR3022 was used as positive control, three different irrelevant human mAbs were used as negative control. **C-D** shows reactivity of historic samples from 50 HIV+ individuals to mRBD and mSpike of SARS-CoV-2. Both HNIG and serum samples from HIV+ donors were collected before the SARS-CoV-2 pandemic.

### The assay can measure antibodies in serum and plasma as well as with and without heat inactivation

One complexity with measuring antibodies in bodily fluids of COVID19 patients is, that infectious virus could be present in the biospecimen, especially early during acute infection. One precaution that is, therefore, often used to limit this risk is to heat inactivate serum or plasma for 1 hour at 56°C. To test if such a heat treatment has an effect on detecting antibodies to the SARS-CoV2 RBD and spike, we compared reactivity of matched non-treated and heat-treated serum samples. While slight differences were observed, they were minimal suggesting that heat treatment has no negative impact on assay performance (**Figure 4A and B**). Similarly, we tested matched serum and plasma samples from patients with COVID19 and found negligible differences suggesting that both types of specimens can be used in the assay interchangeably (**Figure 4C and D**).

**Figure 4:**
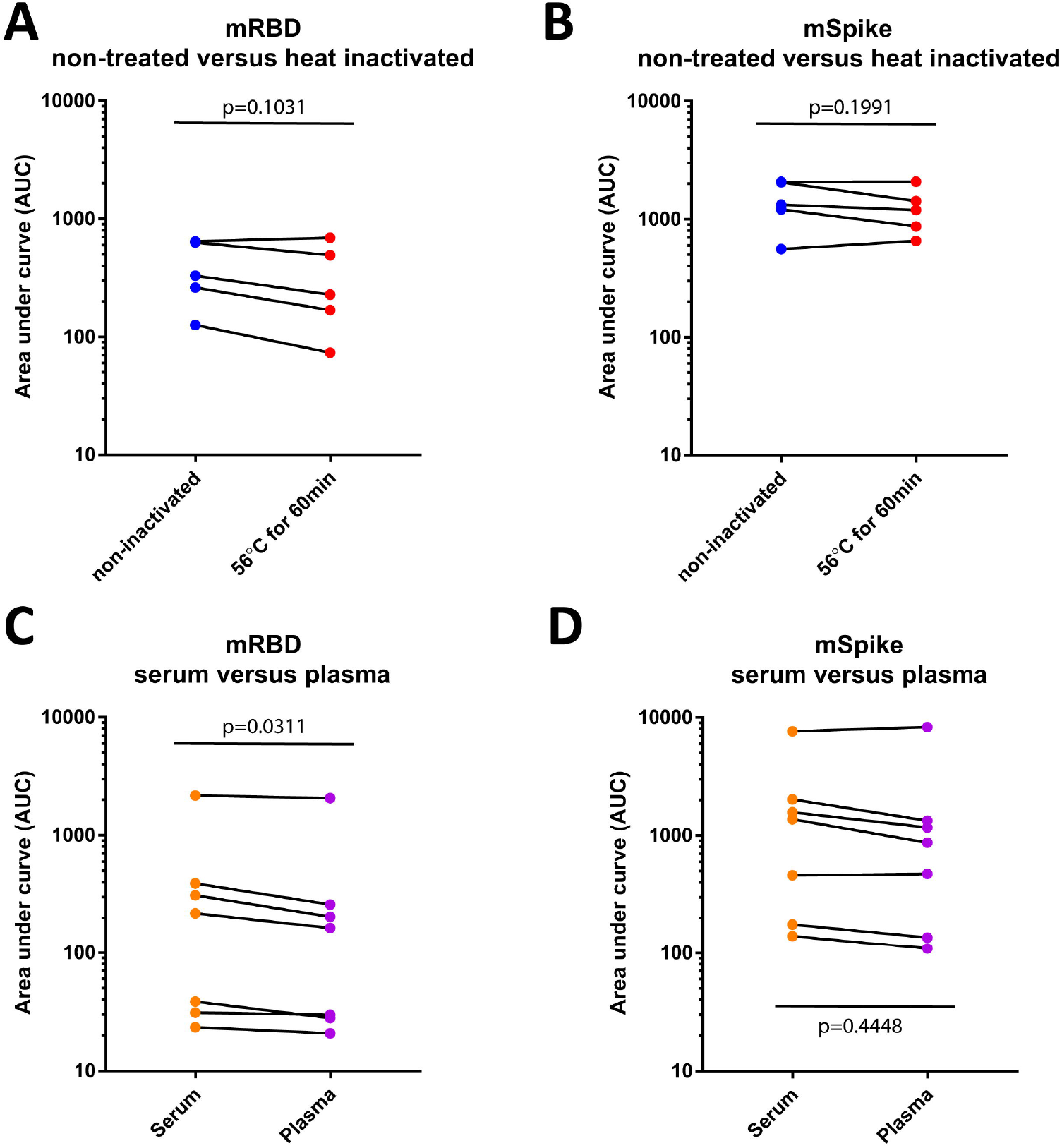
Effect of heat treatment and serum versus plasma on assay performance. **A-B** Reactivity of paired non-treated serum and heat treated serum samples to mRBD and mSpike of SARS-CoV-2 (n=5). **C-D** Reactivity of paired serum and plasma samples to mRBD and mSpike of SARS-CoV-2 (n=7). Statistics were performed using a paired student’s t-test in Graphpad Prism.

### Antibody isotyping, subtyping and neutralizing activity

For the four COVID19 patient plasma/sera from our initial panel, we performed an isotyping and subtyping ELISA using the mammalian cell expressed S proteins. Strong reactivity was found for all samples for IgG3, IgM and IgA (**Figure 5A)**. An IgG1 signal was detected for the majority of samples; low reactivity for IgG2 (in five samples) and IgG4 (in four samples) was also detected. Data from this isotyping ELISA suggest that the IgG response is dominated by IgG3 subtype. Furthermore, we correlated the ELISA reactivity with the neutralizing activity of sera against the USA-WA1/2020 isolate. ELISA titers and microneutralization titers correlated significantly (**Figure 5B)** with a Spearman r of 0.9279.

**Figure 5:**
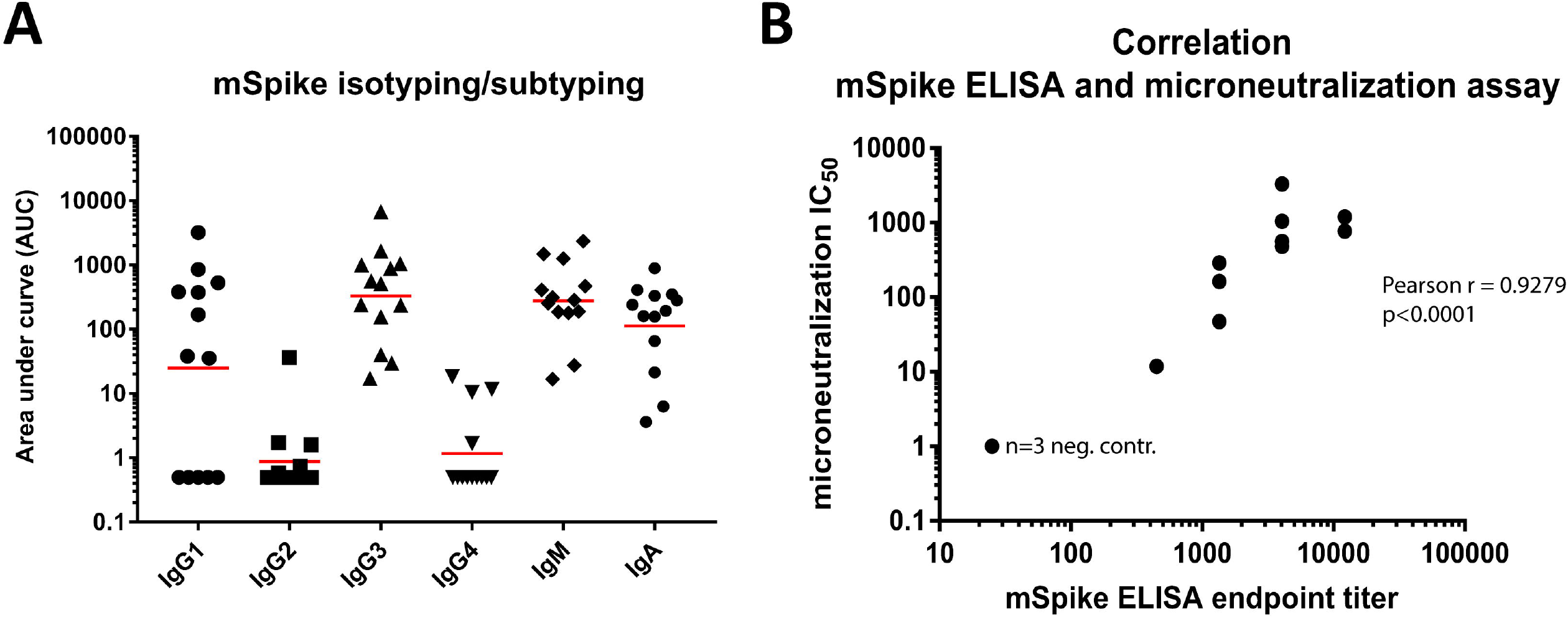
Isotypes and subtypes of antibodies from COVID19 patients to the soluble spike protein and correlation between ELISA and microneutralization titer. **(A)** Mammalian cell derived spike protein was used to study isotype/subclass distribution of antibodies (n=13 positive samples). **(B)** Correlation between ELISA titers and microneutralization titers (n=12, the three samples from negative control sera overlap and are displayed as single point). Statistics were performed using Pearson’s rank test in Graphpad Prism.

## Discussion

Here we describe a serological method to detect seroconversion upon SARS-CoV-2 infection with high specificity and sensitivity. The method is based on reactivity to the immunogenic S protein of the virus. The method is relatively simple and quick in its execution and can be performed at biosafety level 2 level as it does not involve live virus. We have tested these methods using banked serum samples and NHIG preparations obtained from individuals before SARS-CoV-2 started to widely circulate in the US. These serum samples produced low, close to baseline signals in our ELISAs. The age range of the participants was broad, ranging from 20 to 65+ years of age and it is likely that most of these individuals had experienced infections with human coronaviruses including the alphacoronaviruses NL63 and 229E as well as the betacoronaviruses OC43 and HKU1. In fact, the majority of our negative control subjects had strong reactivity to the spike of NL63 and 229E (but showed no cross-reactivity to SARS-CoV-2 RBD and spike). We also included paired serum samples (acute and convalescent) from a participant with a laboratory confirmed coronavirus NL63 infection. Our data show that there is no or only negligible cross-reactivity from human coronaviruses to SARS-CoV-2. Of note, even infection with the human alphacoronavirus NL63, which also uses ACE2 as receptor^23^, did not induce cross-reactivity. Similar findings were reported in a recent preprint where sera from negative control subjects reacted well with spike proteins from human coronavirus but not with SARS-CoV-2.^24^ This is of great importance because it suggests that humans are completely naïve to SARS-CoV-2, which may explain the relatively high R_0_ of SARS-CoV-2 compared to other respiratory viruses such as influenza virus.^25^ It might also suggest that antibody-dependent enhancement from human coronavirus induced cross-reactive antibodies targeted at the S protein is unlikely to be the cause of the high pathogenicity of the virus in humans.^26^

Our data show strong seroconversion after natural infection with SARS-CoV-2. Results from our assays suggest that antibodies mounted upon infection target the full length S protein as well as the RBD, which is the major target for neutralizing antibodies for related viruses coronaviruses.^9^ In fact, sample SARS-CoV2 #1 was tested in another study in neutralization assays and showed a neutralizing titer of 1:160.^13^ In addition, we performed microneutralization assays with a subset of our samples and found excellent correlation between our ELISA titers against the spike protein and virus neutralization. Several of the sera from individuals with confirmed COVID19 showed very strong neutralizing activity with 50% inhibitory concentrations in the hundreds and thousands. Thus, seroconversion might lead to protection from reinfection. Of course, protection – and antibody titers correlated with protection – need to be determined in the near future. In fact, studies to determine whether antibody titers correlate with protection from COVID19 should be one of the highest research priorities at this time. Of note, the ELISA reagents used are derived from the original sequence from Wuhan, the neutralization assays were performed with USA-WA1/2020 (an Asian lineage strain) while the majority of sera were obtained from subjects infected with European-lineage viruses.^27^ The observed correlation between ELISA and neutralization assays hints at minimal antigenic changes. Another interesting finding was, that the IgG3 response appeared stronger than the IgG1 response which is in contrast to e.g. the immune response to influenza where usually IgG1 responses dominate.^28,29^ This is of interest since IgG3 has a stronger affinity to activating Fc-receptors but a shorter half-life than IgG1. We also detected strong IgA and IgM responses in the blood compartment. Of note, level of reactivity and antibody isotypes closely matched expected patterns based on time since symptom onset very well. Antibody isotype and subtype titers were determined using specific secondary antibodies and future studies; however, need to confirm this finding using independent methods.

We also evaluated if heat inactivation can interfere with detection of antibodies. Heat inactivation at 56°C is a standard precaution for work with human sera in many laboratories. A recent pre-print had shown that a commercial AIE/quantum dot-based fluorescence immunochromatographic assay (AFIA, KingFocus Biomedical Engineering Co.,Ltd) assays failed to detect antibodies in samples treated at 56°C for 30 min.^30^ We had made similar observations with a lateral flow assay from BioMedomics. However, our assay was able to detect a signal in samples heat treated for 60 min at 56°C comparable to a signal obtained from non-heat treated sera. Similarly, we found that matched serum and plasma samples showed similar reactivity in the ELISA making it very versatile in terms of which type of specimen can be used.

We did not formally assess specificity and sensitivity of our assay since it might be implemented with different readouts (e.g. OD at a certain dilution, AUC, endpoint titer etc.) and since the assay itself will slightly vary depending on the laboratory that will implement it. However, it is clear from our data that the assay has a high specificity and sensitivity. In addition, our assays are able to measure a quantitative titer which correlates well with neutralization of virus. This is in contrast to many commercial assays that have recently become available on the market which deliver only a qualitative result. Of note, many of these assays have not been independently validated and some of them have been shown to be unfit for the purpose of specifically detecting seroconversion after SARS-CoV-2 infections (e.g. in the UK; Loredo, Texas etc.).

We believe that our ELISA method will be key for serosurveys aimed at determining the real attack rate and infection fatality rate in different human populations and to map the kinetics of the antibody response to SARS-CoV-2. While we found seroconversion in severe, mild and asymptomatic cases, it is possible that some individuals do not seroconvert or that antibody titers wane within short periods of time. To be able to interpret serosurveys correctly, studies to assess the kinetics of the antibody response and the rate of non-responders are urgently needed. Clinical trials with convalescent serum as therapeutic have been initiated in China (e.g. NCT04264858). In addition, a recent report suggests that compassionate use of these interventions could be successful.^31^ Screening potential plasma donors for high antibody titers using our assay is faster and easier than performing standard neutralization assays in BSL3 containment laboratories. Our assay has already been implemented for this purpose in Mount Sinai’s Clinical Laboratory Improvement Amendments (CLIA) regulated clinical laboratory and has received emergency use authorization from New York State. Indeed, more than 20 patients with COVID19 have been compassionately treated at Mount Sinai Hospital with antibody rich plasma from convalescent donors identified with our assays. Last but not least, we believe that our assays could be used to screen health care workers to allow selective deployment of immune medical personnel to care for patients with COVID19. Such a strategy would likely limit nosocomial spread of the virus. More generally, individuals with strong antibody responses could return to normal life, something that might be especially important if a second or third pandemic wave makes quarantine measures again necessary. Importantly, the assumption that individuals with antibodies to the SARS-CoV-2 confer protection from reinfection needs to be confirmed and studies to investigate this should be started as soon as possible. Of course, the generated recombinant proteins are also excellent reagents for vaccine development and can serve as baits for sorting B cells for monoclonal antibody generation. We are making the methods and laboratory reagents widely available to the research community in order to support the global effort to limit and mitigate spread of SARS-CoV-2. A detailed protocol^14^ for antigen expression and ELISA set up is available from the corresponding author and plasmids and proteins have so far been shared with more than 200 laboratories worldwide and also deposited at BEI Resources.

## Methods

### Recombinant proteins

The mammalian cell codon optimized nucleotide sequence coding for the spike protein of SARS-CoV-2 isolate (GenBank: MN908947.3) was synthesized commercially (GeneWiz). The receptor binding domain (RBD, amino acid 319 to 541, RVQP….CVNF) along with the signal peptide (amino acid 1-14, MFVF….TSGS) plus a hexahistidine tag was cloned into mammalian expression vector pCAGGS as well as in a modified pFastBacDual vectors for expression in baculovirus system. The soluble version of the spike protein (amino acids 1-1213, MFVF….IKWP) including a C-terminal thrombin cleavage site, T4 foldon trimerization domain and hexahistidine tag was also cloned into pCAGGS. The protein sequence was modified to remove the polybasic cleavage site (RRAR to A) and two stabilizing mutations were introduced as well (K986P and V987P, wild type numbering). Recombinant proteins were produced using the well-established baculovirus expression system and this system has been published in great detail in ^17,32,33^ including a video guide. Recombinant proteins were also produced in Expi293F cells (ThermoFisher) by transfections of these cells with purified DNA using ExpiFectamine 293 Transfection Kit (ThermoFisher). Supernatants from transfected cells were harvested on day 3 post-transfection by centrifugation of the culture at 4000 g for 20 minutes. Supernatant was then incubated with 6 mls Ni-NTA agarose (Qiagen) for 1-2 hours at room temperature. Next, gravity flow columns were used to collect the Ni-NTA agarose and the protein was eluted. Each protein was concentrated in Amicon centrifugal units (EMD Millipore) and re-suspended in phosphate buffered saline (PBS). Proteins were analyzed on reducing SDS-PAGE. The DNA sequence for all constructs is available from the Krammer laboratory. Several of the expression plasmids and proteins have also been submitted to BEI Resources and can be requested from their web page for free (https://www.beiresources.org/). S1 proteins of NL63 and 229E were obtained from Sino Biologics (produced in 293HEK cells, hexa-histidine tagged). A detailed protocol for protein expression of RBD and spike in mammalian cells is also available.^14^

### SDS-PAGE

Recombinant proteins were analyzed via a standard SDS-PAGE gel to check protein integrity. One ug of protein was mixed with 2X Laemmli buffer containing 5% beta-mercaptoethanol (BME) at a ratio of 1:1. Samples were heated at 100 °Celsius for 15 minutes and then loaded onto a polyacrylamide gel (5% to 20% gradient; Bio-Rad). Gels were stained with SimplyBlue SafeStain (Invitrogen) for 1-2 hours and then de-stained in distilled water overnight.

### Human samples

Human plasma and serum samples were obtained from a number of different sources.

First, de-identified samples from the University of Melbourne (n=3, taken on day 2, 4 and 6 after symptom onset) and University of Helsinki (n=1, day 20 after symptom onset, neutralizing titers 1:160)^13^ were used as positive controls. For those, human experimental work was conducted according to the Declaration of Helsinki Principles and according to the Australian National Health and Medical Research Council Code of Practice. All participants provided written informed consent prior to the study. The studies were approved by the Alfred Hospital (ID #280/14) and University of Melbourne (ID #1442952.1, 1955465.2) Human Research Ethics Committees, and under research permit for project TYH2018322 of Helsinki University Hospital Laboratory.

Second, banked human samples were collected from study participants enrolled in several ongoing IRB approved longitudinal observational study protocols of the Mount Sinai Personalized Virology Initiative. The pre-pandemic serum panel comprised samples selected based on the date of collection (e.g., fall 2019) and whether participants had a documented history of viral infection (e.g., dengue virus, hantavirus, Chikungunya virus, coronavirus NL63). All participants agreed to sample banking and future research use. Self-reported ethnicities of the individuals from which samples were tested included Caucasian, Asian, African American and Hispanic. Samples included sera from a participant with acute NL63 infection as determined by the Biofire Respiratory panel. We included serum collected at day 3 post symptom onset as well as convalescent serum from the same person (day 30 post symptom onset). These samples served as negative controls given that they were collect prior to SARS-CoV-2 spread in the US. Six subjects were 20-29, 19 were 30-39, 13 were 40-49, 7 were 50-59 years old and six were 60 or older. For the mRBD ELISAs sera from additional nine subjects were tested (30-39: 2; 40-49: 4; 50-59: 2; 60+: 1). The pre-pandemic panel was complemented by a collection of plasma samples collected from 50 HIV-1 infected individuals between 2008 and 2011.

The COVID19 panel comprised serum and plasma samples from individuals with severe, mild or asymptomatic SARS-CoV-2 infections. Seven paired serum and plasma samples from patients with COVID19 were used for comparison purposes. These samples were collected between 7 and 31 days post symptom onset.

### Normal human immunoglobulin (NHIG)

The following NHIG preparations, each prepared from >1000 blood/ plasma donors and intended for intravenous use for medical conditions, were tested in an ELISA to determine if they have reactivity against SARS-CoV-2 spike or RBD: Octagam (M934A8541), Gamunex-c (B2GMD00943, A1GLD01882, B3GLD01223, A1GLD01902, B2GLD01972, B3GGD00143, A1GKE00012 (2 different vials), B2GKD00863, B2GJE00033 (3 different vials)), Gammagard liquid (LE12T292AB, LE12V238AB, LE12V278AD), Gammagard S/D (LE08V027AB, 4 different vials), Gammagard liquid (C19G080AAA, LE12V071AD, LE12V230AB, LE12V115AC, LE12V205AB, LE12VE25AB, LE12V115AC).

### ELISA

The ELISA protocol was adapted from previously established protocols ^34,35^. Ninety-six well plates (Immulon 4 HBX; Thermo Scientific) were coated overnight at 4°Celsius with 50 ul per well of a 2 ug/ml solution of each respective protein suspended in PBS (Gibco). The next morning, the coating solution was removed and 100 ul per well of 3% non-fat milk prepared in PBS with 0.1% Tween 20 (TPBS) was added to the plates at room temperature (RT) for 1 hour as blocking solution. Serum samples were heated at 56°C for 1 hour before use to reduce risk from any potential residual virus in serum. Serial dilutions of serum and antibody samples were prepared in 1% non-fat milk prepared in TPBS. The blocking solution was removed and 100 ul of each serial dilution was added to the plates for 2 hours at RT. Next, the plates were washed thrice with 250ul per well of 0.1% TPBS. Next, a 1:3000 dilution of goat anti-human IgG-horseradish peroxidase (HRP) conjugated secondary antibody (ThermoFisher Scientific) was prepared in 0.1% TPBS and 100 ul of this secondary antibody was added to each well for 1 hour. Plates were again washed thrice with 0.1% TBS. Once completely dry, 100 ul of SigmaFast OPD (*o*-phenylenediamine dihydrochloride; Sigma-Aldrich) solution was added to each well. This substrate was left on the plates for 10 minutes and then the reaction was stopped by addition of 50 μL per well of 3 M hydrochloric acid (HCl). The optical density at 490 nanometers was measured via a Synergy 4 (BioTek) plate reader. The background value was set at and optical density (OD) 490nm of 0.11 and area under the curve (AUC) was calculated. AUC values below 1 were assigned a value of 0.5 for graphing and calculation purposes. Data were analyzed using Prism 7 (Graphpad). In some cases endpoint titers were calculated, the endpoint titer being the last dilution before reactivity dropped below and OD 490nm of below 0.11. To determine the impact of heat treatments, paired samples that were heat treated or not treated were analyzed. NHIGs were run similar as serum/plasma samples but with a starting dilution at a concentration of 100 ug/ml. Three non-SARS-CoV-2 reactive human mAbs and CR3022^20-22^, a human mAb reactive to the RBD of both SARS-CoV-1 and SARS-CoV-2 were used as controls. To assess the distribution of the different antibody isotypes/subclasses in the samples that reacted well in our standard ELISA, another ELISA was performed with different secondary antibodies ^29^. These antibodies include anti-human IgA (α-chain-specific) HRP antibody (Sigma A0295) (1:3,000), anti-human IgM (μ-chain-specific) HRP antibody (Sigma A6907) (1:3,000), anti-human IgG1 Fc-HRP (Southern Biotech 9054-05) (1:3,000), anti-human IgG2 Fc-HRP (Southern Biotech #9060-05) (1:3,000), anti-human IgG3hinge-HRP (Southern Biotech 9210-05) (1:3,000), and anti-human IgG4 Fc-HRP (Southern Biotech 9200-05).

### Microneutralization assay

Vero.E6 cells were seeded at a density of 20,000 cells per well in a 96-well cell culture plate in cDMEM. The following day, heat inactivated serum samples (dilution of 1:10) were serially diluted 3-fold in 2X MEM (20% 10× minimal essential medium (Gibco), 4⍰mM L-glutamine, 0.2% of sodium bicarbonate [wt/vol; Gibco], 20⍰mM 4-(2-hydroxyethyl)-1-piperazineethanesulfonic acid (HEPES, Gibco), 200⍰U/ml penicillin–200⍰μ/ml streptomycin (Gibco), and 0.4% bovine serum albumin (MP Biomedical)). The authentic SARS-CoV-2 virus (USA-WA1/2020, GenBank: MT020880) was diluted to a concentration of 100 50% cell culture infectious doses (TCID_50_) in 2xMEM. Eighty μL of each serum dilution and 80μL of the virus dilution were added to a 96-well cell culture plate and allowed to incubate for 1 hr at room temperature. cDMEM was removed from Vero.E6 cells and 120 μL of the virus-serum mixture was added to the cells and the cells were incubated at 37°C for 1 hr. After the 1 hr incubation, the virus-serum mixture was removed from the cells and 100 μL of each corresponding serum dilution and 100 μL of 2X MEM containing 2% FBS (Corning) was added to the cells. The cells were incubated for 48 hr at 37°C and then fixed with 10% paraformaldehyde (PFA) (Polysciences, Inc) for 24 hr at 4°C. Following fixation, the PFA was removed and the cells were washed with 200 μL of PBS. The cells were then permeabilized by the addition of 150 μL of PBS containing 0.1% Triton X-100 for 15 minutes at room temperature. The plates were then washed three times with PBS containing 0.1% Tween 20 (PBS-T) and blocked in blocking solution (3% milk [American Bio] in PBS-T) for 1⍰h at room temperature. After blocking, 100 μL of 1C7 (anti-SARS NP antibody generated in house) at a dilution of 1:1000 was added to all wells and the plates were allowed to incubate for 1 hr at room temperature. Plates were then washed three times with PBS-T before the addition of goat anti-mouse IgG-horseradish peroxidase (IgG-HRP; Rockland Immunochemicals) (diluted 1:3000) in blocking solution for 1⍰hr at room temperature. Plates were then washed three times with PBS-T and the *O*-phenylenediamine dihydrochloride (OPD) substrate (SigmaFast OPD; Sigma-Aldrich) was added. After a 10-minute room temperature incubation, the reaction was stopped by adding 50⍰μL of 3⍰M HCl to the mixture. The optical density (OD) was measured at 490 nm on a Synergy 4 plate reader (BioTek). A cutoff value of the average of the OD values of blank wells plus three standard deviations was established for each plate and used for calculating the microneutralization titer.

### Statistical analysis

Differences between negative controls and positive controls were analyzed using an unpaired t-test. Differences between paired non-treated and heat-treated samples as well as paired serum and plasma samples were analyzed using a paired t-test. Correlation between ELISA titers and neutralization titers were analyzed using Spearman’s rank test. Analyses were performed in GraphPad Prism.

## Data Availability

Data are available from the corresponding author.

## Acknowledgements

We would like to thank Yong-Zhen Zhang (Fudan University) and Eddie Holmes (University of Sydney) for sharing the sequence of the first SARS-CoV-2 isolate in a very timely manner. We thank Jill Garlick and Janine Roney (Alfred Hospital, Melbourne) for data and specimen collection and Nouran Aboelregal for making lots of different NHIGs products (Mount Sinai) available. We are also thankful to Genewiz for speeding up gene synthesis for this project, and being very accommodating to our needs. Furthermore, we want to thank Donna Tidmore for help with ordering primers with near light speed and and finally Susie (Changsu) Dong for commuting to New Jersey on several occasions to pick up reagents from Genewiz. We also thank the study participants for providing biospecimen for research purposes and the Conduits: Mount Sinai Health System Translational Science Hub (NIH grant U54TR001433) for supporting sample collection. The work of the Personalized Virology Initiative is supported by institutional funds and philanthropic donations. This work was partially supported by the NIAID Centers of Excellence for Influenza Research and Surveillance (CEIRS) contract HHSN272201400008C, the Australian National Health and Medical Research Council (NHMRC) NHMRC Program Grant (1071916) and NHMRC Research Fellowship Level B (#1102792), the Academy of Finland and Helsinki University Hospital Funds (TYH2018322). Furthermore, we thank our generous community for providing essential funds and support for our SARS-CoV-2 and COVID-19 research efforts. The following reagent was deposited by the Centers for Disease Control and Prevention and obtained through BEI Resources, NIAID, NIH: SARS-Related Coronavirus 2, Isolate USA-WA1/2020, NR-52281. Finally, we want to thank all the study participants for their contribution to research. We wish the patients with COVID19 a speedy recovery.

## Data availability statement

The data shown in the manuscript is available upon request from the corresponding author.

